# Cell-composition sensitivity of pooled whole-blood DNA methylation profiling in sepsis using the Infinium MethylationEPIC v2.0 array and SeSAMe

**DOI:** 10.64898/2026.05.29.26354469

**Authors:** Piotr K Janicki, Anthony S Bonavia

## Abstract

**Objective:** To characterize pool-level methylation differences associated with sepsis and examine their sensitivity to estimated leukocyte composition using the Infinium MethylationEPIC v2.0 array.

**Design:** Single-center pilot epigenome-wide association study (EWAS) of pooled genomic DNA, analyzed at the pool level with SeSAMe (R/Bioconductor).

**Setting:** Academic medical center.

**Patients:** Fifty critically ill adults enrolled within 48 hours of illness onset and 20 healthy adults. Sepsis was defined by Sepsis-3. All critically ill non-septic patients required mechanical ventilation and/or vasopressors.

**Interventions:** None.

**Measurements and Main Results:** Seventy individual samples were assigned to 14 intended pools of five people (7 sepsis, 3 critically ill non-septic, 4 healthy), of which 13 pools were analyzable: 7 sepsis, 2 critically ill non-sepsis, and 4 healthy. IDATs were processed with SeSAMe QCDPB. Seven blood-cell fractions were estimated with EpiDISH RPC (centDHSbloodDMC). Differential methylation was tested with SeSAMe DML/DMR. Both comparisons used an unadjusted model (∼ Group). The healthy-control adjusted model was ∼ Group + CD8T + CD4T + NK + Mono + B, and the critically ill non-sepsis adjusted model was ∼ Group + CD8T + Mono.

Sepsis pools were neutrophil-dominant (mean Neutro approximately 0.96). Healthy pools had lower neutrophil and higher lymphocyte fractions, whereas the two critically ill non-sepsis pools resembled the sepsis pools. The unadjusted healthy-versus-sepsis DMR analysis produced a very large FDR-significant set, consistent with leukocyte composition rather than many independent locus effects. After multi-probe and effect-size filtering, 86 segments remained. The cell-adjusted model yielded 7 FDR-significant segments with at least 5 CpG. Six adjusted Seg_Est values were positive and the 9q31 interval was negative. None of these segments was recovered in the same direction without adjustment, and unadjusted mean beta at several candidate intervals was higher in healthy pools than in sepsis pools. With four versus seven pools and five fraction covariates, the adjusted model was over-specified and signs could change.

In the critically ill non-sepsis versus sepsis comparison (2 versus 7 pools), the unadjusted analysis yielded 26 FDR-significant segments, of which 10 remained after filtering for at least 5 CpGs and |Seg_Est| ≥ 0.001. The adjusted analysis identified 18 FDR-significant multi-probe segments; 17 estimates were negative and one was positive. PLSCR1, TRIOBP, and PGLYRP1 were highlighted because of their prior relevance to sepsis.

**Conclusions:** The observed methylation differences tracked estimated leukocyte composition and could not be attributed specifically to sepsis. The study demonstrates the technical feasibility of generating EPIC v2 data from pooled DNA, but it does not validate pooling for sepsis EWAS or establish reproducible sepsis-specific DMRs. The findings are exploratory because inference was conducted at the pool level, comparator groups differed substantially, adjusted models were underpowered, and individual-level and sorted-cell validation were unavailable.

## INTRODUCTION

Sepsis is characterized by a complex and temporally dynamic immune response in which inflammatory and immunosuppressive programs coexist rather than evolve in a simple linear sequence [1, 2]. This dysregulated host response is increasingly understood to involve persistent reprogramming of innate immune cells, particularly within the myeloid compartment. Monocytes and macrophages may acquire durable phenotypic changes after pathogen exposure, resulting in tolerance or trained immunity that shapes subsequent responses to infection [3]. The mechanisms that stabilize these altered immune states, however, remain incompletely defined.

Epigenetic regulation is a plausible mediator of this process. DNA methylation can influence chromatin accessibility and transcription factor binding, thereby linking inflammatory stimuli to sustained changes in gene expression. In myeloid cells, methylation dynamics at promoter and enhancer regions regulate lineage-defining transcriptional programs, including those governed by PU.1 (SPI1) and C/EBPalpha (CEBPA) [4, 5]. Experimental models have shown that trained immunity and tolerance are accompanied by coordinated epigenetic remodeling, including DNA methylation changes at regulatory loci [3]. Clinical studies extend these observations by demonstrating that methylation patterns in circulating immune cells are associated with inflammatory signaling and organ dysfunction in sepsis [6].

Epigenome-wide studies in critically ill patients have reported methylation differences associated with sepsis. Prior analyses have identified differentially methylated regions enriched in innate immune pathways, including genes involved in complement activation, endothelial dysfunction, and antimicrobial responses [7, 8]. Beltran-Garcia et al. reported that leukocyte methylation changes in sepsis and septic shock were associated with severe clinical phenotypes and enrichment of pathways linked to immunosuppression [8]. These studies have also suggested a predominance of global hypomethylation in sepsis, consistent with coordinated activation of host-defense programs [9]. However, the current literature remains limited by small cohorts, methodological heterogeneity, and reliance on earlier array platforms with incomplete coverage of distal regulatory elements.

The Infinium Methylation EPIC v2.0 array offers broader interrogation of the methylome, including enhancers, super-enhancers, and noncoding regulatory regions relevant to immune-cell biology [10]. At the same time, large-scale epigenomic studies in sepsis remain constrained by cost, DNA input requirements, and sample availability, particularly in biobank settings. DNA pooling has emerged as a practical strategy to reduce these barriers, and prior work suggests that pooled methylation profiling can preserve biologically meaningful signals under selected conditions [11]. Its performance in sepsis, however, where inter-individual heterogeneity is substantial, remains uncertain.

We therefore characterized genome-wide differential methylation in a pilot critical-care cohort using the EPIC v2.0 platform and pooled whole-blood DNA. We also examined how estimated leukocyte composition affected pool-level regional methylation results.

## METHODS

### Patient Recruitment and Sample Pooling

This prospective, single-center pilot study enrolled critically ill adults (≥18 years) within 48 hours of illness onset at an academic medical center. Patients were initially identified using an electronic medical record-based screening strategy and subsequently confirmed by clinician review. Eligibility required organ support, defined as mechanical ventilation and/or vasopressor therapy. Sepsis was defined according to Sepsis-3 criteria [12], requiring suspected or confirmed infection with an increase in Sequential Organ Failure Assessment (SOFA) score of ≥2 points. A total of 50 critically ill patients were enrolled under IRB #15328. In addition, 20 healthy adult controls provided samples through a commercial genomic DNA company (Polbiomed LLC) with explicit consent for collection and use of de-identified data.

Control samples were derived from two comparator subgroups. The first subgroup comprised critically ill adults requiring organ support who did not meet Sepsis-3 criteria and had no suspected or confirmed infection at enrollment. The second subgroup comprised healthy adult controls. For methylation analysis, 70 individual samples were organized into 14 intended pools of five individuals each, comprising 7 sepsis pools (n = 35), 3 critically ill non-septic pools (n = 15), and 4 healthy-control pools (n = 20). Pool assignment used a weighted Gower-like distance based on sepsis status, age, and sarcopenia, followed by greedy nearest-neighbor grouping [13]. Pool-level age, sex distribution, and mean SOFA values for the critically ill pooled subset are summarized in Supplementary Tables S1 and S2.

Clinical blood was collected in EDTA tubes and stored as whole blood without prior freeze-thaw. DNA was isolated using the Kurabo QuickGene DNA Whole Blood system. Five contributor aliquots were combined per pool using equal intended DNA mass. Healthy-control and clinical specimens were processed asynchronously.

One critically ill non-sepsis pool was excluded because of an excessive number of missing methylation measurements, consistent with poor DNA extraction or quality. The final analytic dataset comprised 13 pools: 7 sepsis pools, 2 critically ill non-sepsis pools, and 4 healthy-control pools. Because the two comparator groups differed clinically and in estimated leukocyte composition, analyses were performed separately for sepsis versus healthy pools and sepsis versus critically ill non-sepsis pools. Both comparisons were considered exploratory.

### Bioinformatic analysis

Whole-blood DNA methylation was measured on the Infinium MethylationEPIC v2.0 array (∼937 000 loci). IDAT files were processed in SeSAMe with the QCDPB pipeline. Probes with ≥20% missing values and probes on the recommended EPICv2 mask were removed. Blood cell-type fractions (B, NK, CD4T, CD8T, monocytes, neutrophils, eosinophils) were estimated with EpiDISH robust partial correlations (RPC) [14–17] using the centDHSbloodDMC reference after collapsing replicate probe prefixes. Differential methylation was tested at the locus and segment levels with SeSAMe DML/DMR (EPICv2, hg38). Both comparisons used an unadjusted model (∼ Group), with the relevant non-sepsis group as the reference. The healthy-control adjusted model was ∼ Group + CD8T + CD4T + NK + Mono + B. The critically ill non-sepsis adjusted model was ∼ Group + CD8T + Mono because B-cell, NK-cell, and CD4 T-cell fractions were estimated as zero in all nine pools. Neutrophil and eosinophil fractions were omitted to avoid including all components of a closed composition. For descriptive summaries of the unadjusted regional catalogues, segments were required to have segment-level FDR < 0.05, at least 5 CpGs, and |Seg_Est| ≥ 0.10 for healthy controls versus sepsis or |Seg_Est| ≥ 0.001 for critically ill non-sepsis versus sepsis. Estimates with |Seg_Est| ≥ 1 were excluded. Given the small number of pools, the unadjusted regional catalogues were interpreted descriptively rather than as evidence of independent locus-level discoveries.

### Raw β display of adjusted segments

For segments identified by the adjusted models, unadjusted mean β values were plotted by pool for the relevant comparison. Agreement between the unadjusted group difference (sepsis minus comparator) and adjusted Seg_Est was used only as a descriptive check. Opposite signs indicate that the adjusted coefficient is conditional on estimated cell composition and is not reflected in the unadjusted β distribution.

### Software used

R version 4.5.3 (2026-03-11 ucrt), SeSAMe 1.28.1, Bioconductor 3.22, EpiDish 2.26.0.

## RESULTS

### Deconvolution of study pools

Table 1 summarizes EpiDISH RPC fractions estimated with the centDHSbloodDMC reference. Sepsis pools (SEP-P01 to SEP-P07) were strongly neutrophil-dominant (mean estimated neutrophil fraction 0.96, range 0.94-0.97), with B-cell, NK-cell, CD4 T-cell, and eosinophil fractions estimated as zero. Critically ill non-sepsis pools (CINS-P01 and CINS-P02) resembled the sepsis pools. Healthy pools (HC-P01 to HC-P04) had lower estimated neutrophil fractions (0.67-0.84) and detectable lymphocyte fractions. Combining the two comparator groups would therefore mix distinct estimated leukocyte compositions.

**Table 1.** EpiDISH RPC estimated cell fractions. Rows sum to approximately 1. CINS-P01 and CINS-P02 are critically ill non-sepsis pools; HC-P01 to HC-P04 are healthy pools; and SEP-P01 to SEP-P07 are sepsis pools. P values are from descriptive two-sided Welch tests comparing all non-sepsis pools with sepsis pools. Eosinophil estimates were zero in every pool, so no P value was calculated.

|  | **B** | **NK** | **CD4T** | **CD8T** | **Mono** | **Neutro** | **Eosino** |
| --- | --- | --- | --- | --- | --- | --- | --- |
| CINS-P01 | 0 | 0 | 0 | 0.011 | 0.057 | 0.931 | 0 |
| CINS-P02 | 0 | 0 | 0 | 0.016 | 0.012 | 0.972 | 0 |
| HC-P01 | 0.005 | 0.023 | 0.019 | 0.066 | 0.042 | 0.844 | 0 |
| HC-P02 | 0.033 | 0.051 | 0.079 | 0.117 | 0.055 | 0.665 | 0 |
| HC-P03 | 0.02 | 0.028 | 0.057 | 0.057 | 0.085 | 0.754 | 0 |
| HC-P04 | 0.019 | 0.078 | 0.007 | 0.105 | 0.061 | 0.73 | 0 |
| mean | **0.013** | **0.030** | **0.027** | **0.062** | **0.052** | **0.816** | 0.000 |
| SD | 0.013 | 0.030 | 0.033 | 0.044 | 0.024 | 0.120 | 0.000 |
| SEP-P01 | 0 | 0 | 0 | 0 | 0.029 | 0.971 | 0 |
| SEP-P02 | 0 | 0 | 0 | 0.007 | 0.029 | 0.965 | 0 |
| SEP-P03 | 0 | 0 | 0 | 0.021 | 0.042 | 0.937 | 0 |
| SEP-P04 | 0 | 0 | 0 | 0.006 | 0.027 | 0.968 | 0 |
| SEP-P05 | 0 | 0 | 0 | 0.003 | 0.032 | 0.965 | 0 |
| SEP-P06 | 0 | 0 | 0 | 0.024 | 0.036 | 0.94 | 0 |
| SEP-P07 | 0 | 0 | 0 | 0.017 | 0.043 | 0.94 | 0 |
| mean | 0.000 | 0.000 | 0.000 | 0.011 | 0.034 | 0.955 | 0.000 |
| SD | 0.000 | 0.000 | 0.000 | 0.009 | 0.006 | 0.015 | 0.000 |
| P | 0.065 | 0.06 | 0.103 | 0.038 | 0.13 | 0.036 |  |

In descriptive two-sided Welch tests comparing all six non-sepsis pools with sepsis pools, P values were 0.038 for CD8 T cells and 0.036 for neutrophils; B-cell and NK-cell comparisons yielded P values of 0.065 and 0.060. The six comparisons were not independent because the estimated fractions are compositional and several were structurally zero. They were not interpreted as independent discoveries.

Because group differences in estimated leukocyte composition could produce apparent regional methylation differences, cell fractions were included as covariates in adjusted analyses. Healthy and critically ill non-sepsis pools were analyzed separately against the sepsis pools.

### DMR analysis (healthy controls vs sepsis)

The unadjusted SeSAMe DMR analysis (β ∼ Group) yielded 187,992 segments with segment-level FDR < 0.05. This large catalogue coincided with the marked difference in estimated leukocyte composition and should not be interpreted as 187,992 independent sepsis loci. After descriptive filtering for at least 5 CpGs and |Seg_Est| ≥ 0.10, 86 autosomal segments remained (Supplementary Table S3): 64 had lower and 22 had higher estimated methylation in sepsis relative to healthy pools.

Cell-type adjustment used EpiDISH RPC fractions and the model

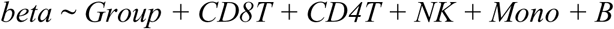

Neutrophil and eosinophil fractions were omitted so that the design did not include all components of a closed composition. The reported condition number of the non-zero fraction matrix was 21,901, indicating severe collinearity and unstable adjusted coefficient estimates.

Seven adjusted segments met FDR < 0.05 and contained at least five CpGs (Table 2). Six had positive Seg_Est values (+0.18 to +0.79), and one interval at 9q31 had Seg_Est = –0.31. No autosomal segment from Table 2 was recovered in the same direction in both models after the same multi-probe and effect-size filter. At the adjusted intervals in Table 2, unadjusted mean beta was higher in healthy pools than in sepsis pools, opposite to several positive adjusted coefficients (Figure 1). The adjusted sign is therefore a composition-conditioned residual rather than a raw beta contrast.

**Table 2.** Adjusted DMRs in healthy-control pools versus sepsis pools, restricted to segments with at least five CpGs (FDR < 0.05). Seg_Est is the Group coefficient with healthy control as the reference; positive values indicate higher adjusted methylation in sepsis.

| Seg_ID | hg38 | bp | n | Seg_Est | FDR | Positional annotation |
| --- | --- | --- | --- | --- | --- | --- |
| 141609 | chr6:29,553,368–29,554,005 | 638 | 21 | +0.35 | $2.05 \times 10^{-12}$ | MHC class I region |
| 17431 | chr1:56,645,049–56,645,839 | 791 | 14 | +0.22 | $1.16 \times 10^{-4}$ | *PLPP3* |
| 211277 | chr9:113,463,504–113,464,730 | 1227 | 7 | −0.31 | $5.07 \times 10^{-4}$ | 9q31; SVEP1/LPAR1 |
| 222157 | chr10:18,140,105–18,140,832 | 728 | 9 | +0.28 | $1.77 \times 10^{-2}$ | 10p12 (CACNB2) |
| 247637 | chr11:40,293,068–40,294,312 | 1245 | 9 | +0.43 | $4.28 \times 10^{-2}$ | 11p12 |
| 37241 | chr1:224,929,687–224,930,116 | 430 | 8 | +0.31 | $4.26 \times 10^{-2}$ | 1q42 |
| 132879 | chr5:169,300,748–169,301,210 | 463 | 8 | +0.54 | $4.28 \times 10^{-2}$ | *DOCK2* |

These data show a large, composition-linked methylation difference between the healthy and sepsis pools. They do not define a reproducible set of sepsis-specific DMRs. Neighborhood labels in Table 2 are positional and do not establish altered gene function.

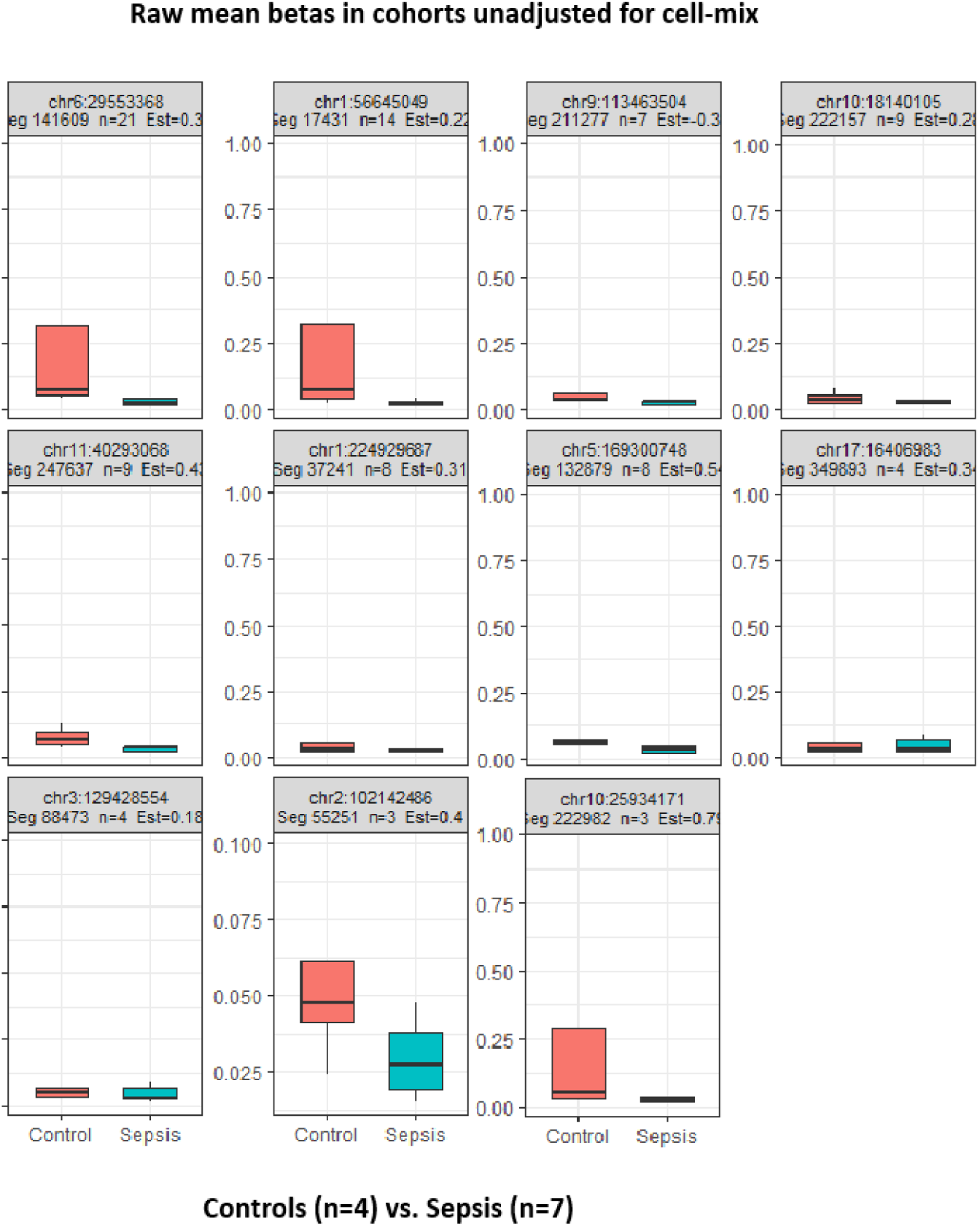

### DMR analysis (critically ill vs sepsis)

The unadjusted SeSAMe DMR analysis (β ∼ Group) yielded 26 autosomal segments with segment-level FDR < 0.05 (Supplementary Table S3), consistent with the smaller estimated composition gap between two neutrophil-rich groups. Descriptive filtering for at least 5 CpGs and |Seg_Est| ≥ 0.001 left 10 segments, all with lower estimated methylation in sepsis than in critically ill non-sepsis pools. This lower effect-size threshold was used because the critically ill comparison did not yield regions meeting the 0.10 threshold used for the healthy-control comparison.

The composition-adjusted model was

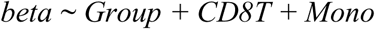

Neutrophil and eosinophil fractions were omitted. In these nine pools, B-cell, NK-cell, and CD4 T-cell fractions were estimated as zero, so only CD8T and monocyte fractions in the model varied. The reported condition number of the non-zero fraction matrix was 101, indicating that adjusted estimates remained sensitive to collinearity.

Table 3 lists 18 FDR-significant multi-probe segments from the adjusted analysis, with critically ill non-sepsis pools as the reference group. Ten of the 18 adjusted intervals overlapped the 10 filtered unadjusted DMRs and had the same direction of effect. PLSCR1, TRIOBP, and PGLYRP1 are highlighted because of their prior relevance to sepsis.

**Table 3.** Adjusted DMRs, critically ill pools (n = 2) vs sepsis pools (n = 7). Seg_Est = Group coefficient (reference = critically ill).

| Seg_ID | chr | start | end | Seg_Est | n | SYMBOL | FDR |
| --- | --- | --- | --- | --- | --- | --- | --- |
| 383275 | chr19 | 9674835 | 9675456 | -0.05074 | 12 | ZNF562 | 1.78E-13 |
| **90801** | **chr3** | **146544387** | **1.47E+08** | **-0.02961** | **11** | **PLSCR1** | **3.25E-11** |
| **421849** | **chr22** | **37696644** | **37697073** | **-0.0383** | **10** | **TRIOBP** | **2.65E-10** |
| 145548 | chr6 | 32177369 | 32178819 | -0.01295 | 33 | AGPAT1 | 5.41E-07 |
| 122626 | chr5 | 69332604 | 69332912 | -0.05571 | 6 | CCDC125 | 2.35E-05 |
| 280509 | chr12 | 85836026 | 85836603 | -0.06273 | 5 | RASSF9 | 2.69E-05 |
| 53639 | chr2 | 88170211 | 88170655 | -0.05242 | 5 | THNSL2 | 3.87E-05 |
| 107898 | chr4 | 98143171 | 98143754 | -0.02562 | 9 | STPG2 | 0.000104 |
| 350379 | chr17 | 5500571 | 5501262 | -0.03415 | 8 | SMIM48 | 0.000149 |
| 313955 | chr14 | 104085530 | 1.04E+08 | -0.04303 | 6 | ASPG | 0.000371 |
| 65010 | chr2 | 197786156 | 1.98E+08 | -0.04712 | 9 | BOLL | 0.001459 |
| 125011 | chr5 | 93620768 | 93621695 | -0.01492 | 10 | MIR2277 | 0.005228 |
| **392972** | **chr19** | **46023063** | **46023493** | **-0.02574** | **9** | **PGLYRP1** | **0.008365** |
| 166648 | chr7 | 19772856 | 19772970 | -0.01795 | 5 | TMEM196 | 0.010807 |
| 315227 | chr15 | 23565702 | 23567188 | 0.075462 | 5 | MKRN3 | 0.022704 |
| 329283 | chr15 | 101127938 | 1.01E+08 | -0.06069 | 5 | LOC124903564 | 0.03262 |
| 191991 | chr8 | 43140421 | 43141492 | -0.0265 | 5 | HGSNAT | 0.046547 |
| 13193 | chr1 | 38872764 | 38873882 | -0.00193 | 13 | RRAGC-DT | 0.046547 |

Seventeen of the 18 Seg_Est values in Table 3 were negative, corresponding to lower adjusted methylation estimates in sepsis; the MKRN3 interval was positive. The raw beta distributions for the three highlighted intervals were directionally consistent with the adjusted estimates, but this concordance does not establish independence from cell composition. The comparison included only two critically ill non-sepsis pools, and several estimated fractions were structurally zero.

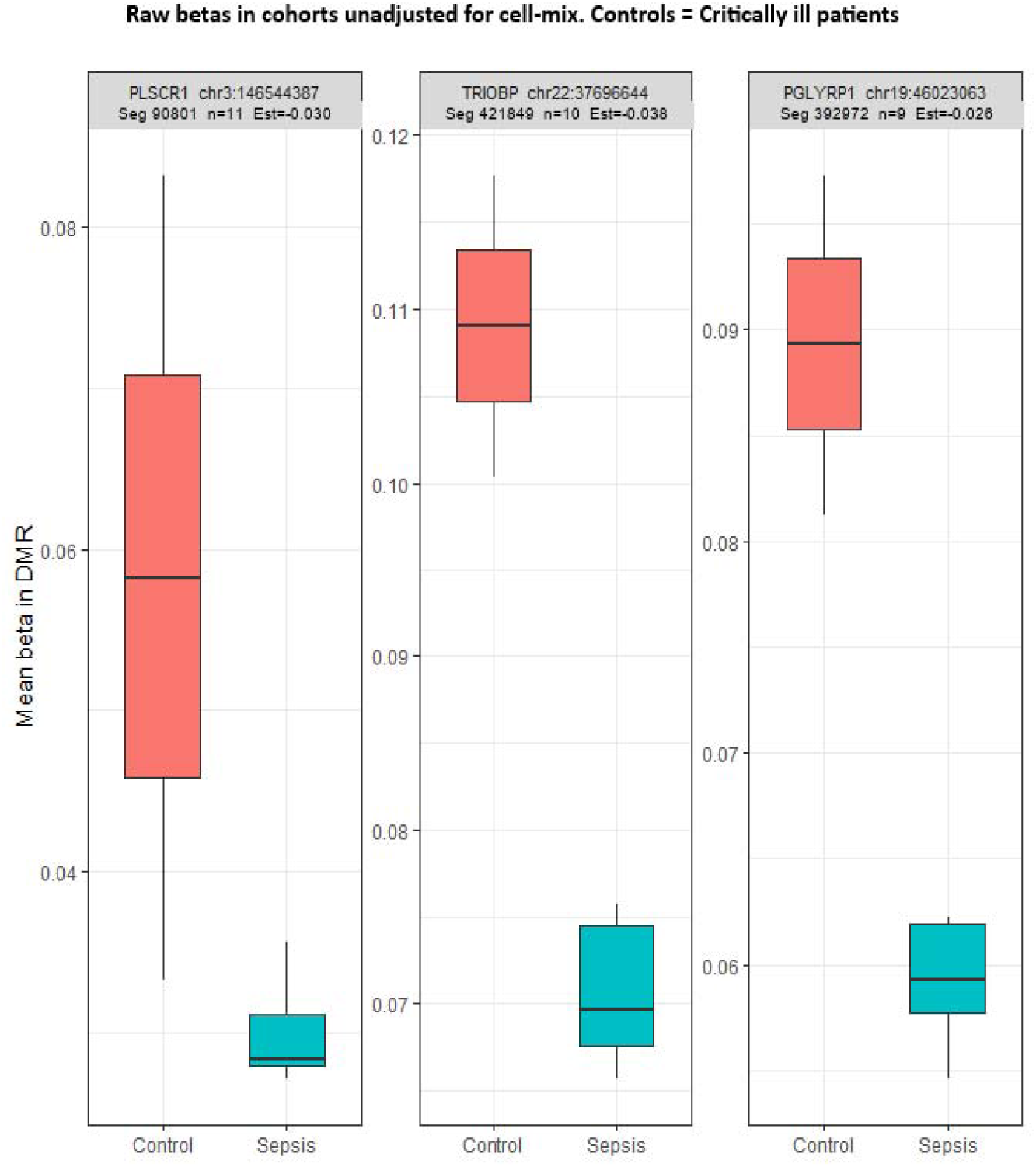

## DISCUSSION

The principal finding was that the two comparator groups had distinct estimated leukocyte compositions. Sepsis pools were strongly neutrophil-dominant, as were the two critically ill non-sepsis pools. Healthy pools had lower estimated neutrophil fractions and detectable lymphocyte fractions. The regional analyses were therefore performed separately for healthy versus sepsis pools and critically ill non-sepsis versus sepsis pools.

EPIC v2 whole-blood profiles differ between sepsis and comparator pools, but that difference is not separable from leukocyte composition at this sample size. Unadjusted DMR (∼ Group) in healthy vs sepsis produced an extremely large number of FDR-significant segments, the expected footprint of Neutro ≈ 0.82 vs ≈ 0.96 rather than of many independent locus effects. After multi-probe and effect-size filters, 86 segments remained (64 hypo– and 22 hypermethylated in sepsis). No overlapping autosomal segments were recovered in both the unadjusted and cell-adjusted models.

The composition-adjusted model yielded 11 segments with FDR < 0.05 and at least three CpGs. Seven contained at least five CpGs and are presented in Table 2. Ten of the 11 had positive Seg_Est values (+0.18 to +0.79); the interval at 9q31 had Seg_Est = –0.31. These intervals lie near MHC class I, *PLPP3*, *DOCK2*, and a 9q31 neighborhood that includes *SVEP1* and *LPAR1* [20]. That neighborhood is of interest because of prior genetic findings, not because this study measured those genes or variants. Nakada et al. reported that *SVEP1* c.2080A>C (p.Gln581His, rs10817033) was associated with 28-day mortality and organ failure in European-ancestry septic shock, and that *SVEP1* silencing increased LPS-driven endothelial IL-8, GRO-alpha, MCP-1, and MCP-3 [19]. *LPAR1*, adjacent at 9q31.3, encodes an LPA receptor involved in leukocyte migration and endothelial barrier function. These reports support listing 9q31 as a positional annotation. They do not show that the present DMR overlaps the coding SNP, alters *SVEP1*/*LPAR1* transcription, or that study participants carry rs10817033. Genotyping was not performed, so the methylation results do not support recommending SNP panels.

The same limit applies to MHC class I. HLA molecules are central to antigen presentation and have been linked to sepsis risk in candidate studies (including a reported association of HLA-A*31 with sepsis susceptibility) [20, 21]. Whole-blood methylation near MHC is also highly sensitive to cell mix and haplotype structure. Unadjusted mean β at several Table 2 intervals was higher in controls than in sepsis, whereas several adjusted Seg_Est values were positive, the opposite of a naive reading of the adjusted coefficient. With four healthy pools versus seven sepsis pools and five fraction covariates, the healthy-control adjusted model was over-specified, and coefficient signs could change. Adjustment-specific DMRs should be reported as composition-conditioned residuals, not as disease-specific hypomethylation.

The critically ill non-sepsis versus sepsis contrast more directly addresses sepsis because both groups were neutrophil-dominant. The unadjusted analysis yielded 26 segments, 10 of which remained after the stated multi-probe and effect-size filters. The adjusted analysis yielded 18 FDR-significant multi-probe segments; 17 estimates were negative and one was positive. Ten adjusted intervals overlapped the filtered unadjusted list with the same direction of effect. Three highlighted intervals were annotated to the promoters of *PLSCR1*, *TRIOBP*, and *PGLYRP1* (Table 3). Their adjusted effect estimates were small, approximately 2.6-3.8 percentage points on the β scale, and negative. Because the comparator comprised only two pools and the estimated fractions remained collinear, these intervals should be treated as exploratory positional annotations.

### Limitations

All inference was conducted at the pool level: seven sepsis pools were compared with four healthy pools or two critically ill non-sepsis pools. The small number of pools limited power and prevented estimation of participant-level variability or outcome associations. Age, sex, and severity of illness were not included in the regional models. Healthy-control and clinical samples were processed asynchronously, so specimen source and processing batch are inseparable from the healthy-versus-sepsis comparison. The design was cross-sectional, and individual-level, functional, and sorted-cell validation were unavailable. Residual myeloid confounding also remains possible despite composition adjustment.

The most consistent finding was compositional: sepsis and critically ill non-sepsis pools were neutrophil-dominant, whereas healthy pools were not. The remaining multi-probe intervals are suitable for hypothesis generation, but they do not establish coordinated, sepsis-specific epigenetic remodeling or support clinical stratification or therapeutic targeting.

Larger studies will need an explicitly defined critically ill non-infectious comparator, individual rather than exclusively pooled arrays, a prespecified DMR caller with genome-wide error control (for example, limma with DMRcate or an equivalent approach), and, if 9q31 or MHC remains of interest, paired genotyping and sorted-cell or single-cell methylation rather than SNP recommendations inferred from a positional label.

### Supplemental Tables

**Supplementary Table S1.**
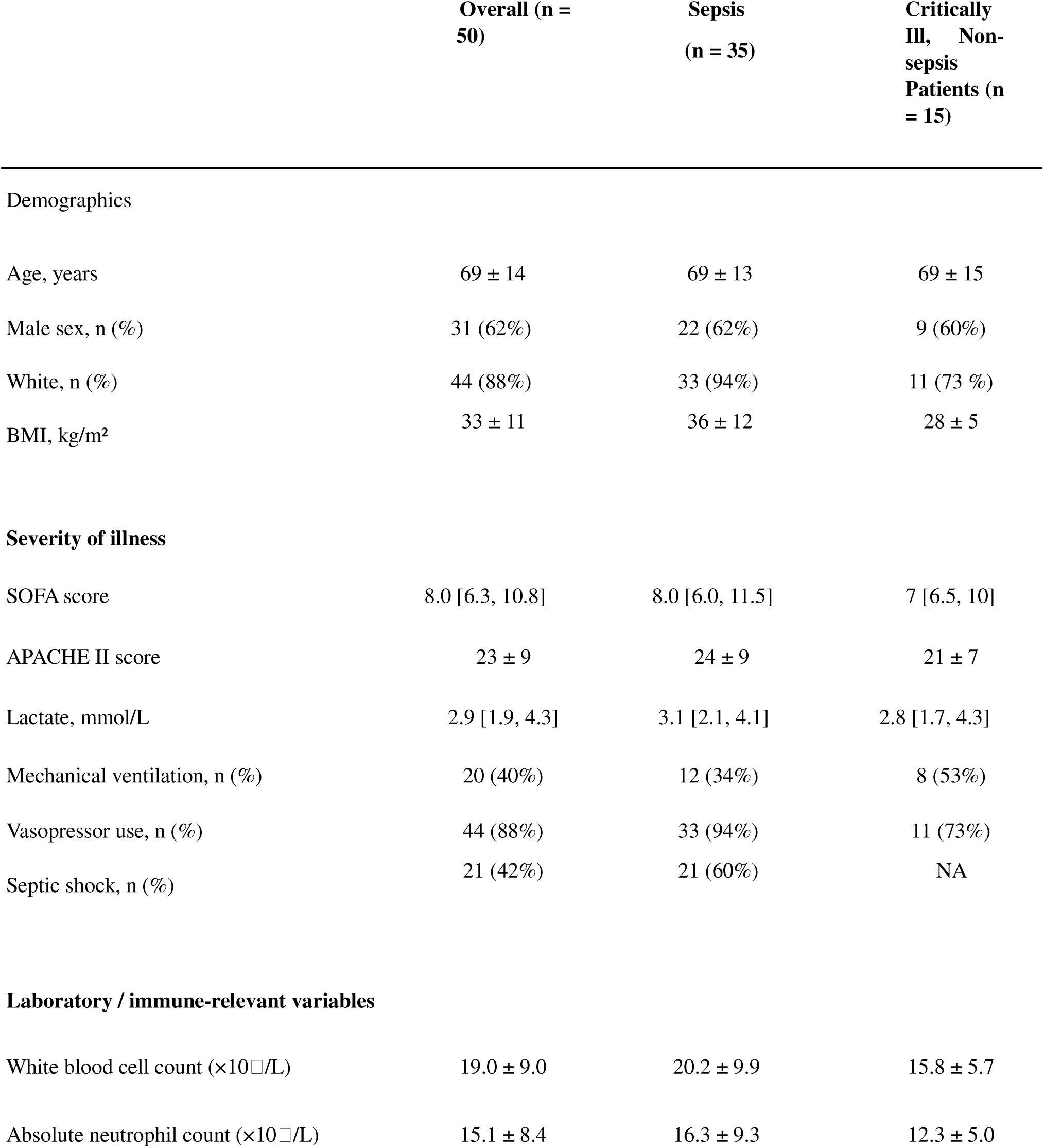
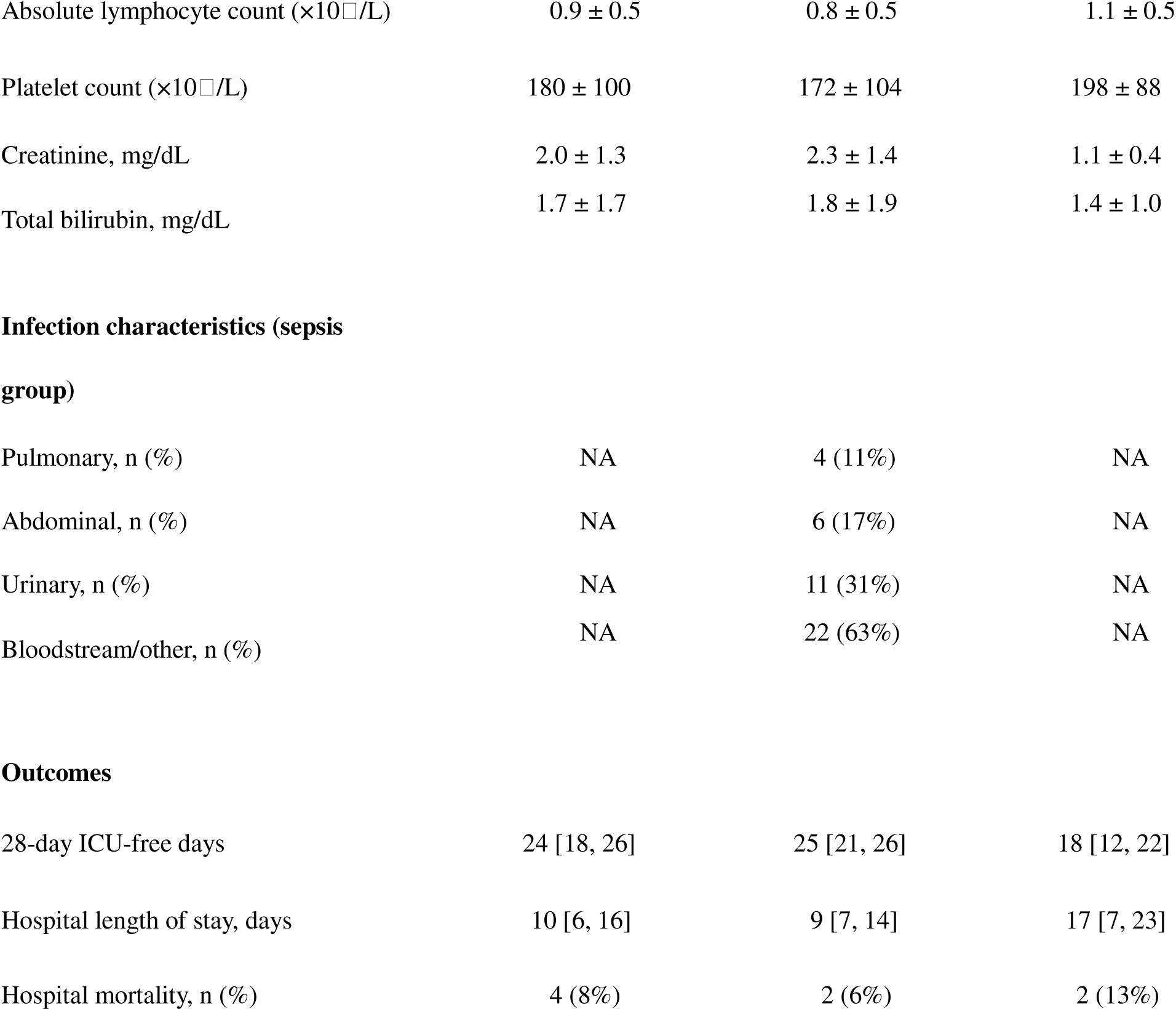
Baseline characteristics of critically ill participants assigned to intended DNA pools.

**Supplementary Table S2.**
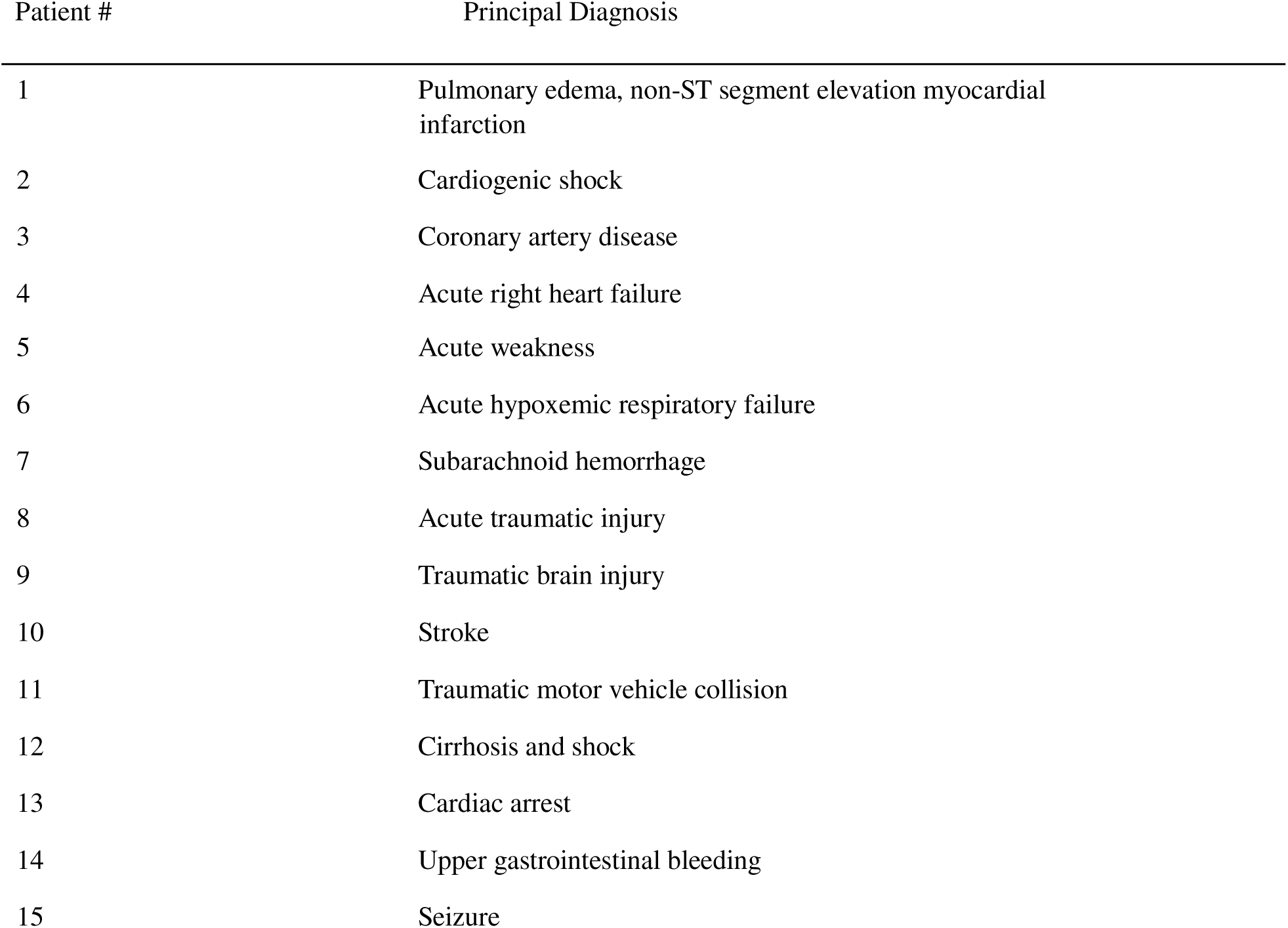
Principal diagnoses in the critically ill non-sepsis cohort.

## DECLARATIONS

### Author Contribution Statement

PKJ and ASB contributed to the conception/design of the study and contributed to data acquisition. PKJ performed data analysis. PKJ and ASB performed data interpretation, drafted the manuscript, and critically revised the manuscript for important intellectual content. All authors approved the final version of the manuscript and agree to be accountable for all aspects of the work.

### Ethics

This study was conducted in accordance with the principles of the Declaration of Helsinki and was approved by the Institutional Review Board (IRB #15328) of the participating academic medical center. For critically ill patients, eligibility screening was performed using electronic medical records and confirmed by clinician review. Given the acute nature of critical illness, informed consent was obtained from patients or their legally authorized representatives, as appropriate.

Samples from healthy control participants were obtained through a commercial genomic DNA assay conducted by Polbiomed LLC. All control participants provided explicit written informed consent for the collection of biological specimens and the use of their de-identified genomic data for research purposes.

All data and biospecimens were de-identified prior to analysis to ensure participant confidentiality.

## Data Availability

All data produced in the present study are available upon reasonable request to the authors

## Notes

### Competing Interest Statement

PKJ is the founder and CEO of Polbiomed LLC (PA, USA).

### Author Declarations

Ethics committee/IRB of Penn State University gave ethical approval for this work

### Summary of Updates

This revision substantially reframes the study to address the major limitations of the original pooled analysis. The objective was changed from validating pooled methylation profiling and identifying a sepsis signature to characterizing pool level methylation differences and determining their sensitivity to estimated leukocyte composition. Healthy controls and critically ill patients without sepsis are no longer combined into one comparator group. They are analyzed separately because they differ clinically and have markedly different estimated blood cell distributions. EpiDISH deconvolution was added for B cells, natural killer cells, CD4 T cells, CD8 T cells, monocytes, neutrophils, and eosinophils. SeSAMe analyses now include both unadjusted and cell composition adjusted models. The adjusted model for the critically ill comparison includes only CD8 T cells and monocytes because several other fractions were estimated as zero in every pool. The preprocessing, quality control, software versions, model specifications, filtering criteria, and sample exclusion are now described more explicitly. Results were recalculated and reconciled across the text, tables, supplementary material, and figures. In the healthy control comparison, the large unadjusted regional catalogue was reduced to 86 regions after descriptive filtering, while the adjusted analysis identified seven regions containing at least five CpG sites. These findings are interpreted primarily as evidence of cell composition effects. For the critically ill comparison, 26 unadjusted regions were identified, 10 remained after descriptive filtering, and 18 adjusted regions were identified. Ten adjusted regions overlapped the filtered unadjusted set with the same direction of effect. PLSCR1, TRIOBP, and PGLYRP1 are presented as exploratory positional findings. The broad pathway, microRNA, and biologic claims derived from the combined comparator analysis were removed. The conclusions now emphasize leukocyte composition, model instability, small numbers of pools, asynchronous sample processing, and the absence of individual level or sorted cell validation. The manuscript therefore presents the findings as exploratory and does not claim a reproducible sepsis specific methylation signature or validation of pooling.

### Funding

- National Institutes of Health: R35GM150695

## REFERENCES

[1] Cao M, Wang G, Xie J. Immune dysregulation in sepsis: experiences, lessons and perspectives. Cell Death Discov 2023;9(1):465.

[2] Wu D, Shi Y, Zhang H, Miao C. Epigenetic mechanisms of Immune remodeling in sepsis: targeting histone modification. Cell Death Dis 2023;14(2):112.

[3] Saeed S, Quintin J, Kerstens HH, Rao NA, Aghajanirefah A, Matarese F, et al. Epigenetic programming of monocyte-to-macrophage differentiation and trained innate immunity. Science 2014;345(6204):1251086.

[4] Netea MG, Quintin J, van der Meer JW. Trained immunity: a memory for innate host defense. Cell Host Microbe 2011;9(5):355–61.

[5] Gosselin D, Link VM, Romanoski CE, Fonseca GJ, Eichenfield DZ, Spann NJ, et al. Environment drives selection and function of enhancers controlling tissue-specific macrophage identities. Cell 2014;159(6):1327–40.

[6] Lorente-Sorolla C, Garcia-Gomez A, Catala-Moll F, Toledano V, Ciudad L, Avendano-Ortiz J, et al. Inflammatory cytokines and organ dysfunction associate with the aberrant DNA methylome of monocytes in sepsis. Genome Med 2019;11(1):66.

[7] Binnie A, Walsh CJ, Hu P, Dwivedi DJ, Fox-Robichaud A, Liaw PC, et al. Epigenetic Profiling in Severe Sepsis: A Pilot Study of DNA Methylation Profiles in Critical Illness. Crit Care Med 2020;48(2):142–50.

[8] Beltran-Garcia J, Casabo-Valles G, Osca-Verdegal R, Navarrete-Lopez P, Rodriguez-Gimillo M, Nacher-Sendra E, et al. Alterations in leukocyte DNA methylome are associated to immunosuppression in severe clinical phenotypes of septic patients. Front Immunol 2023;14:1333705.

[9] Caldwell BA, Wu Y, Wang J, Li L. Altered DNA methylation underlies monocyte dysregulation and immune exhaustion memory in sepsis. Cell Rep 2024;43(3):113894.

[10] Peters TJ, Meyer B, Ryan L, Achinger-Kawecka J, Song J, Campbell EM, et al. Characterisation and reproducibility of the HumanMethylationEPIC v2.0 BeadChip for DNA methylation profiling. BMC Genomics 2024;25(1):251.

[11] Lopez-Cruz I, Garcia-Gimenez JL, Madrazo M, Garcia-Guallarte J, Piles L, Pallardo FV, et al. Epigenome-wide DNA methylation profiling in septic and non-septic patients with similar infections: potential use as sepsis biomarkers. Front Cell Infect Microbiol 2024;14:1532417.

[12] Singer M, Deutschman CS, Seymour CW, Shankar-Hari M, Annane D, Bauer M, et al. The Third International Consensus Definitions for Sepsis and Septic Shock (Sepsis-3). JAMA 2016;315(8):801–10.

[13] Kaplow IM, MacIsaac JL, Mah SM, McEwen LM, Kobor MS, Fraser HB. A pooling-based approach to mapping genetic variants associated with DNA methylation. Genome Res 2015;25(6):907–17.

[14] Teschendorff AE, Breeze CE, Zheng SC, Beck S. A comparison of reference-based algorithms for correcting cell-type heterogeneity in Epigenome-Wide Association Studies. BMC Bioinformatics 2017;18(1):105.

[15] Teschendorff AE, Zheng SC. Cell-type deconvolution in epigenome-wide association studies: a review and recommendations. Epigenomics 2017;9(5):757–68.

[16] Zheng SC, Breeze CE, Beck S, Teschendorff AE. Identification of differentially methylated cell types in epigenome-wide association studies. Nat Methods 2018;15(12):1059–66.

[17] Zheng SC, Webster AP, Dong D, Feber A, Graham DG, Sullivan R, et al. A novel cell-type deconvolution algorithm reveals substantial contamination by immune cells in saliva, buccal and cervix. Epigenomics 2018;10(7):925–40.

[18] Elenbaas JS, Pudupakkam U, Ashworth KJ, Kang CJ, Patel V, Santana K, et al. SVEP1 is an endogenous ligand for the orphan receptor PEAR1. Nat Commun 2023;14(1):850.

[19] Nakada TA, Russell JA, Boyd JH, Thair SA, Walley KR. Identification of a nonsynonymous polymorphism in the SVEP1 gene associated with altered clinical outcomes in septic shock. Crit Care Med 2015;43(1):101–8.

[20] da Silva FP, Preuhs Filho G, Finger E, Barbeiro HV, Zampieri FG, Goulart AC, et al. HLA-A*31 as a marker of genetic susceptibility to sepsis. Rev Bras Ter Intensiva 2013;25(4):284–9.

[21] Guo L, Shen S, Rowley JW, Tolley ND, Jia W, Manne BK, et al. Platelet MHC class I mediates CD8+ T-cell suppression during sepsis. Blood 2021;138(5):401–16.

